# Prevalence of Long COVID in *Mycobacterium tuberculosis-*exposed Groups in Peru and Kenya

**DOI:** 10.1101/2025.04.28.25326537

**Authors:** Asiko Ongaya, Ariana R. Cardenas, Clement Shiluli, Lourdes B. Ramos, Liz C. Senador, Juan A. Flores, Bernard N. Kanoi, Josephine F. Reijneveld, Angel Ruvalcaba, Danny Perez, Paul Waiganjo, Cecilia S. Lindestam Arlehamn, Timothy J. Henrich, Michael J. Peluso, Segundo R. Leon, Jesse Gitaka, Sara Suliman

## Abstract

**Background:** Long COVID (LC), also referred to as post-COVID condition, refers to new or worsening symptoms lasting more than three months after SARS-CoV-2 infection. The prevalence of LC, and the impact of co-infection with prevalent pathogens such as *Mycobacterium tuberculosis* (*Mtb*), in low- and middle-income countries remain unclear. We aimed to address these gaps in two *Mtb*-exposed populations.

**Methods:** We recruited HIV-uninfected pulmonary tuberculosis (TB) patients (n=36) and their household contacts (n=63) in Peru, and healthcare workers (n=202) in Kenya. We collected clinical data using study instruments adapted from a United States based study of LC. Participants were sampled within 2 years of SARS-CoV-2 diagnosis.

**Results:** In Peru, 41.4% participants reported LC symptoms, with no TB-associated significant differences in the prevalence or clinical phenotypes of LC. The most common LC symptoms were neurological (e.g., headache and trouble sleeping) and musculoskeletal (e.g., back pain). Kenyan participants reported acute, but no LC symptoms, and reported a decline in the quality of life during acute infection. In Peru, the post-COVID-19 period was associated with a significant decline in all quality-of-life dimensions (p<0.01), except depression and anxiety (p=0.289).

**Conclusion:** This study shows that LC prevalence was high in Peru, where TB status was not linked to LC symptoms. Those with LC reported high levels of musculoskeletal and neurological symptoms. Unexpectedly, healthcare workers in Kenya denied the presence of LC symptoms. These findings highlight the need for long-term follow-up and larger studies in different geographic settings to dissect the impact of TB comorbidity on LC.

## Introduction

Long COVID (LC) is defined as the persistence or development of new or worsening symptoms that continue to be present at least 3 months after infection with severe acute respiratory syndrome coronavirus 2 (SARS-CoV-2) (1). Intensive efforts are now underway to identify the etiology of this condition (2). Possible contributors include: persistent reservoirs of SARS-CoV-2 (3,4), tissue inflammation, immune dysregulation, microbiota disruption, clotting and endothelial abnormalities, and dysfunctional neurological signaling (5,6). Prior work has identified host-related risk factors for developing LC, which include the female sex (7,8), and the presence of certain comorbidities such as obesity, diabetes, and the presence or reactivation of chronic viral infections (9–11).

LC has been identified in almost all regions of the world affected by COVID-19 (5). Concerns have been raised about its potential impact in low- and middle-income countries (LMICs) where the condition has generally received less attention. To date, there is limited data available from low- and middle-income countries over 90% of LC studies have been conducted in high and upper-middle-income countries (12). Furthermore, key questions remain about the impact of other co-infections which may be vastly more prevalent in LMICs, in particular *Mycobacterium tuberculosis* (*Mtb*) infection, which is the causative agent of Tuberculosis (TB) disease, which is estimated to impact 10 million people annually. Mouse model studies show that preexisting *Mtb* infection decreased SARS-CoV-2 viral loads and weight loss, suggesting a paradoxical protective role of *Mtb* against SARS-CoV-2 (13,14). However, more studies are urgently needed to define the interaction between *Mtb* exposure and LC risk, and to evaluate the mechanisms by which *Mtb* may alter post-acute outcomes of COVID-19.

In this study, we evaluated the prevalence of LC in cohorts from two settings with high burdens of TB (Peru and Kenya) (15). Our study population included healthcare workers (HCWs) (16,17) and household contacts (HHCs) of persons with TB disease (18,19, 20), both of which are at high risk of exposure to *Mtb*. Using the Long-term Impact of Infection with Novel Coronavirus (LIINC) study instruments that have been used to study the natural history of LC in the U.S. (21), we evaluated two previously understudied populations in the Global South to define the prevalence of LC in these *Mtb-*exposed groups.

## Materials and Methods

### Ethics review

This study was approved by Mount Kenya Ethics (MKU/ERC/2252) in Kenya, San Juan Bautista Private University Ethics Committee (N°705-2022-CIEI-UPSJB) in Peru, and the University of California, San Francisco (IRB no. 22-37759). All participants signed informed consent for the study. We conducted the study in accordance with Good Clinical Practice guidelines. HIV infection was an exclusion criterion in both Peru and Kenya.

### Study Design and Participants

#### Lima, Peru

We recruited 99 adult participants from January 2023 to May 2024, which included 36 persons with pulmonary TB and their 63 household contacts (HHCs), which were stratified by the QuantiFERON-TB-Gold plus (QFT) test as described below. All participants had a documented history of previous SARS-CoV-2 infection. They were recruited from 35 community health centers and Maria Auxiliadora Hospital in Southern Lima, Peru. COVID-19 history was determined either by a positive molecular or antigen test or cohabitation with a person with confirmed COVID-19 and developing symptoms within two weeks of exposure. Patients with active TB were diagnosed either by a positive sputum smear or clinical criteria according to the Peruvian National TB guidelines (20). All persons with TB undergoing treatment were invited to participate unless they had a confirmed diagnosis of HIV or any form of drug-resistant TB, which we obtained from their treatment cards and clinical records. To recruit HHCs, we contacted the index cases and conducted a household visit to invite the contacts to participate in the study. HHCs were defined as persons living with the TB patient in the same housing unit. We conducted household visits in Peru at recruitment, as well as two and four months after the initial visit.

#### Nairobi, Kenya

We enrolled 202 participants from Mbagathi Hospital in Nairobi, Kenya. We recruited healthcare workers, including medical doctors, nurses, clinical officers, pharmacists, pharmaceutical technologists, radiologists, laboratory technologists, and technicians, among others, in a cross-sectional study design from April 2023 to July 2024. We collected data and samples at a single visit. We recorded demographic data using a standardized questionnaire by interviewing participants. Clinical measures were taken by a study-trained clinician. We collected blood samples for QFT analysis.

#### Data collection

In both cohorts, we collected epidemiological, clinical, and COVID-19 symptomatology information using an instrument adapted from the UCSF-based Long-term Impact of Infection with Novel Coronavirus (LIINC) study (21) which has been used to assess the presence and severity of post-acute COVID-attributed symptoms and quality of life in San Francisco and has supported phenotyping for multiple studies of Long COVID pathobiology (22,23). The questionnaires were administered by trained interviewers. We collected demographic data including age, sex, education level, and residence type (permanent houses made out of bricks or stones, or non-permanent houses made of cardboard, mud, iron sheets, or wood). Clinical data included body mass index (BMI), and comorbidities, including autoimmune diseases, cancer, diabetes, hypertension, history of heart attack, and other lung conditions (e.g., asthma). We also collected behavioral data, including smoking habits. Alcohol use data were collected according to the WHO-approved test: Development of the Alcohol Use Disorders Identification Test (AUDIT) (24). Each participant was asked for the date of SARS-CoV-2 symptom onset, the date of their SARS-CoV-2 positive test, and the method of diagnosis: PCR or antigen test. Interviews were conducted to gather information on the existence, duration (in days), and present condition of predefined LC symptoms adapted from the LIINC questionnaire (21). If the person had more than one reported SARS-CoV-2 infection episode, we asked for the symptoms present during the most recent infection. Finally, we recorded whether participants had been hospitalized (defined as admitted for more than 24 hours) and if they required supplemental oxygen, admission to an intensive care unit (ICU), or mechanical ventilation. Past medical history and medication intake were also documented.

#### Quality of Life

Quality of life was measured using questions adapted from the EuroQoL metrics (21). The metrics measured 5 domains: mobility, self-care, usual activities, pain, and depression and anxiety, where 1 corresponded to no problems while performing the activity, to 5 corresponding to extreme problems or inability to perform the activity. For the Kenyan Cohort, we reported quality of life scores for three periods: before COVID-19, during COVID-19, and after the acute phase of COVID-19 (i.e. during the time of recruitment for this study). For the Peruvian Cohort, we only recorded quality of life scores, before and after COVID-19, but not during the acute phase of SARS-CoV-2 infection. Self-reported health scores ranged from 0 to 100 on a visual-analog scale.

#### QuantiFERON-TB-Gold Plus (QFT) analysis

We collected whole blood samples from participants at Mbagathi Hospital and health centers in Southern Lima in 6 mL lithium heparin vacuum tubes and transported them within 4 hours of collection at room temperature to the Kenya Medical Research Institute (KEMRI-CRDR) TB laboratory in Kenya, and San Juan Bautista University in Peru for processing. QFT tests were performed to calculate IFN-γ concentrations (IU/mL) following the manufacturer’s instructions. Indeterminate QFT results were repeated, and if they remained indeterminate, they were excluded from the analysis (25).

#### Statistical analysis

We described the categorical variables as frequency rates and percentages, and continuous variables as medians and interquartile ranges (IQRs). We calculated the association between categorical variables and TB groups using Fisher’s exact test. For continuous variables, we applied the Kruskal-Wallis test, with a significance threshold level of 0.05. To report the symptoms, we considered the self-reported presence or absence of the symptom. LC was defined as having at least one COVID-related symptom present at least 90 days following SARS-CoV-2 infection, consistent with the WHO and National Academies of Sciences, Engineering, and Medicine (NASEM) (26,27). If the symptoms persisted at the time of the interview, we calculated the duration by subtracting the symptom onset date from the interview date. Similarly to other studies, we grouped the symptoms into 8 categories according to the impacted body system: cardiopulmonary, constitutional, dermatologic, gastrointestinal, genitourinary, musculoskeletal, neurologic, and upper respiratory (28). Participants were considered within the category if they had at least one symptom from that category.

We calculated the prevalence ratio (PR) using generalized linear models (GLM) with a log link and Poisson robust error variance. Poisson regression analysis for developing LC based on symptoms in the acute phase of 10 days since diagnosis or symptom onset (29). Crude rates were calculated in the univariate model; for the multivariate analysis for each symptom, we included age, sex, and BMI a priori, based on epidemiological criteria (7,30,31). Quality of life questions were reported as frequency rates and percentages. In all TB groups, we compared the quality of life in the period before and after COVID-19, using Fisher’s exact test. Quality of life scores were treated as continuous variables. The statistical analysis was generated with STATA Software (v.17), and figures were built under R software (v.4.3.0) and Prism GraphPad (v.10.3.1).

## Results

### Study participants

We enrolled 99 participants in Peru and 202 participants in Kenya (Table 1). In the Peruvian cohort, there were 36 participants with TB disease, 32 QFT+, and 31 QFT-household contacts (HHC) (Table 1). In this cohort, there were more males among TB participants than the HHCs (27.8%, p<0.001). We asked participants about their alcohol and smoking habits. The majority (87.9%) had a low risk of developing alcohol addiction according to the AUDIT guidelines (24) with no significant differences in the alcohol addiction risk nor smoking habits between the groups. The BMI between the Peruvian participant groups was significantly different (<0.001), with the active TB group having the lowest BMI, as expected (15). There were no significant differences in COVID-19 vaccine doses and the prevalence of comorbidities amongst our cohorts. The most prevalent comorbidities were diabetes, hypertension, and other lung conditions like asthma (Table 1). Due to the limited availability of COVID-19 tests in Peru, we diagnosed COVID-19 either by a confirmed positive antigen or molecular test, or by the development of symptoms within two weeks following exposure to an index SARS-CoV-2-positive case. Participants with TB disease were more likely to have a history of COVID-19 due to proximity to an index COVID-19 case than the household contacts (p=0.029) (Table 1).

**Table 1:**
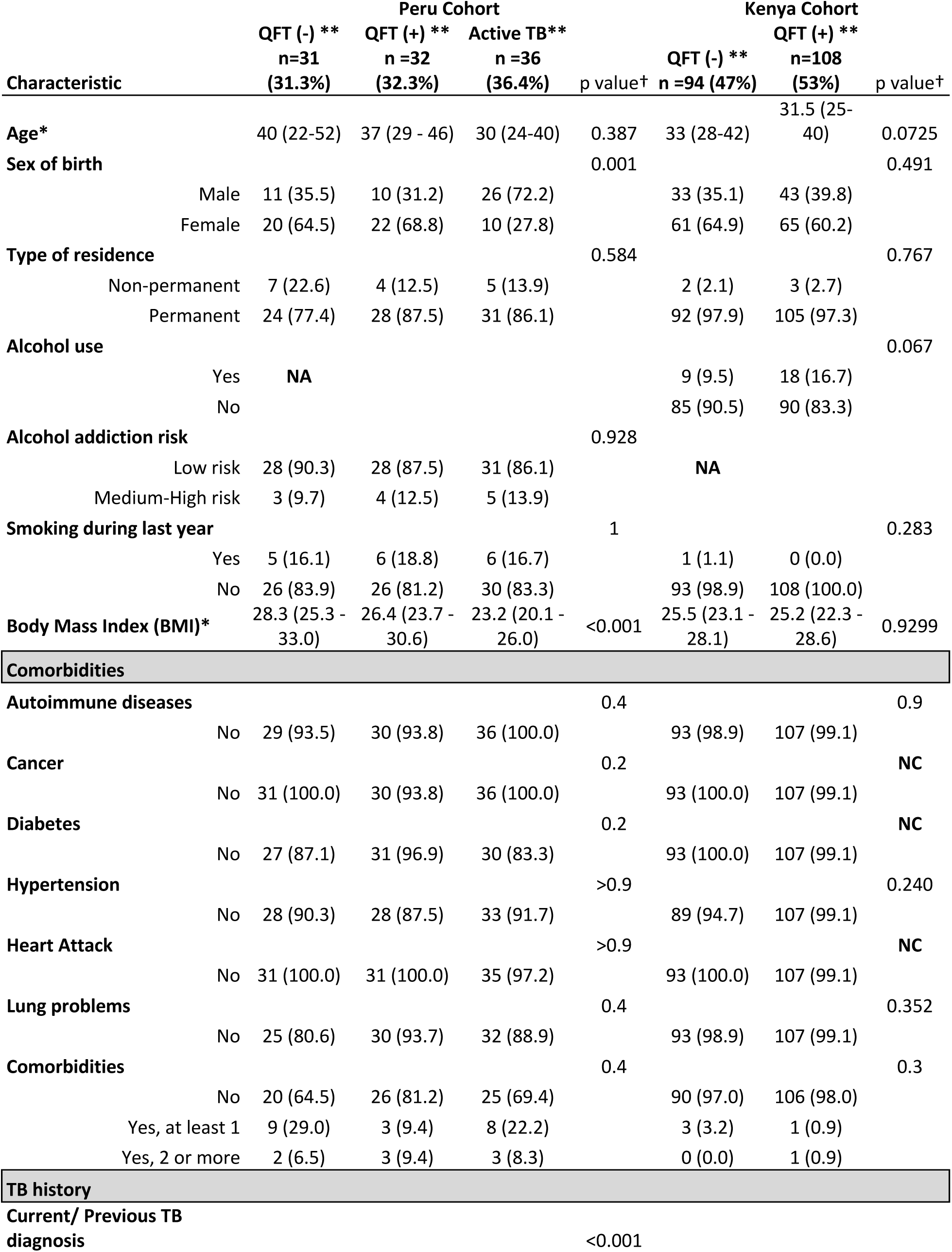
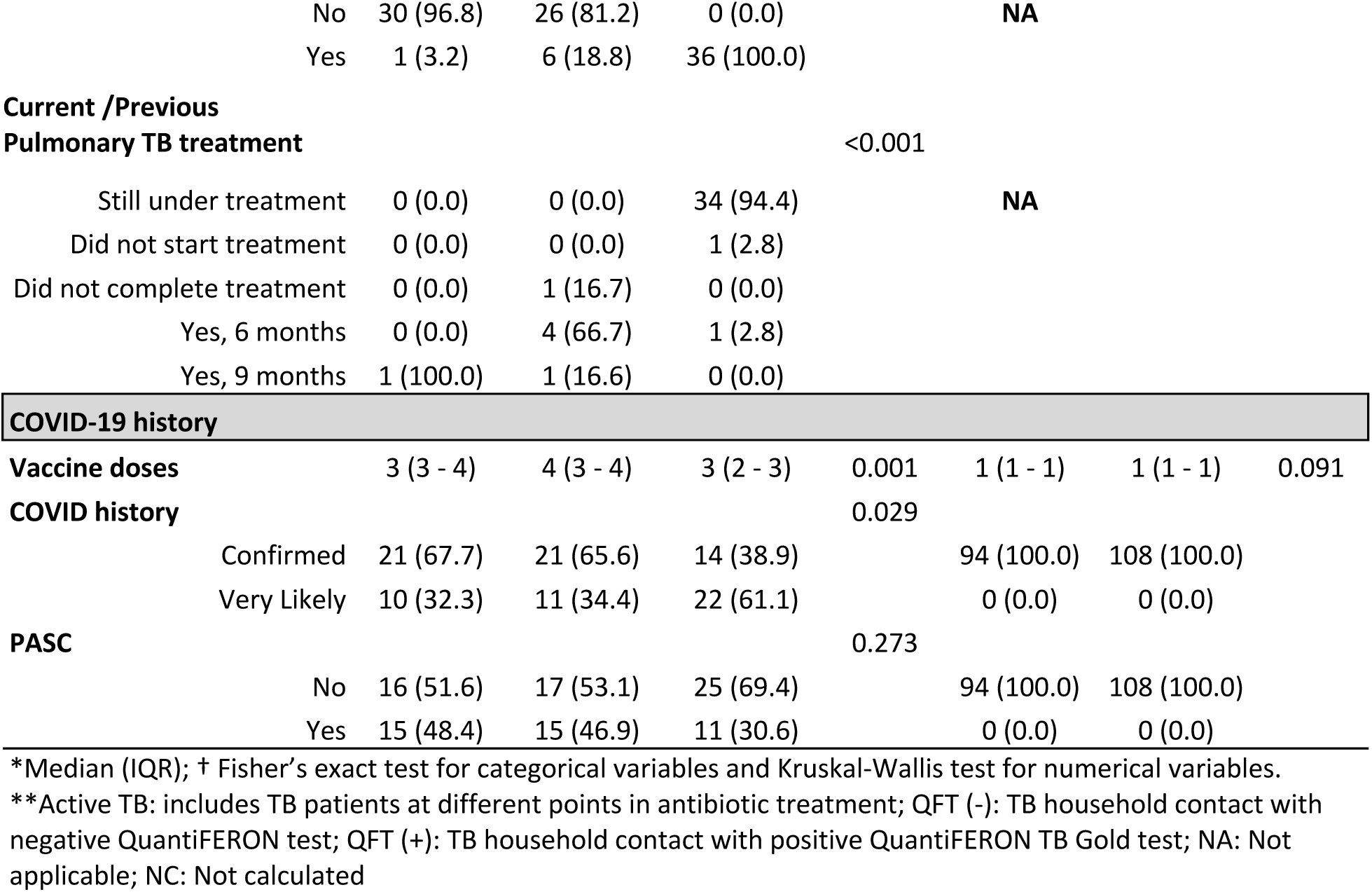
Demographic and clinical data and COVID-19 history.

In the Kenyan cohort, there were 94 QFT-, and 108 QFT+ participants. The median age between groups was similar (Table 1), and both groups had a female majority, with no difference between the two groups. Similarly, alcohol use, smoking habits, and comorbidities were infrequent in the Kenyan cohort. All Kenyan participants received a single COVID-19 vaccine dose. All 202 participants in the Kenyan cohort were diagnosed with COVID-19 by PCR (Table 1).

### Acute COVID-19 Symptoms

In the Peruvian cohort, the most prevalent symptoms of acute COVID-19 were headache (75.8%), tiredness (73.7%), and feeling feverish (69.7%), while in the Kenyan cohort tiredness (60.2%), feeling feverish (55.1%) and chills (50.5%) were prevalent (Table S1).

### Post-acute COVID Symptoms

In the Peruvian cohort, 41.4% of individuals met the criteria for LC. The most prevalent LC symptoms in this cohort (n=99) were back pain (19%), pain in arms and legs (15%) and cough (14%). In contrast to the Peruvian cohort, no individuals in the Kenyan cohort met criteria for LC (Table 1). We also analyzed the durability of COVID-19 symptoms in Kenya, where the median did not exceed 8 days across the different symptoms (Figure 2). In the acute phase, the most durable symptoms were cough and numbness in the QFT- and QFT+ groups, respectively.

### Influence of *Mtb* Infection and TB Disease on Long COVID

We analyzed symptoms meeting the LC definition (>90 days) in the Peruvian cohort, which showed that diarrhea was the most durable symptom in the QFT-group (median [IQR]: 1054 [940-1168] days) and tiredness (861 [580-1124] days). In the QFT+ group, the most common symptoms were menstrual pain (856 [743–969] days) and cough (812 [588-854] days). Finally, in the TB patients in Peru, trouble concentrating was the most durable symptom (1482 days), followed by dizziness, but only in a single participant (1028 days) (Figure 1a). There were no statistical differences in LC prevalence between the groups (p=0.273), suggesting that LC was present in all the groups irrespective of TB status.

**Figure 1:**
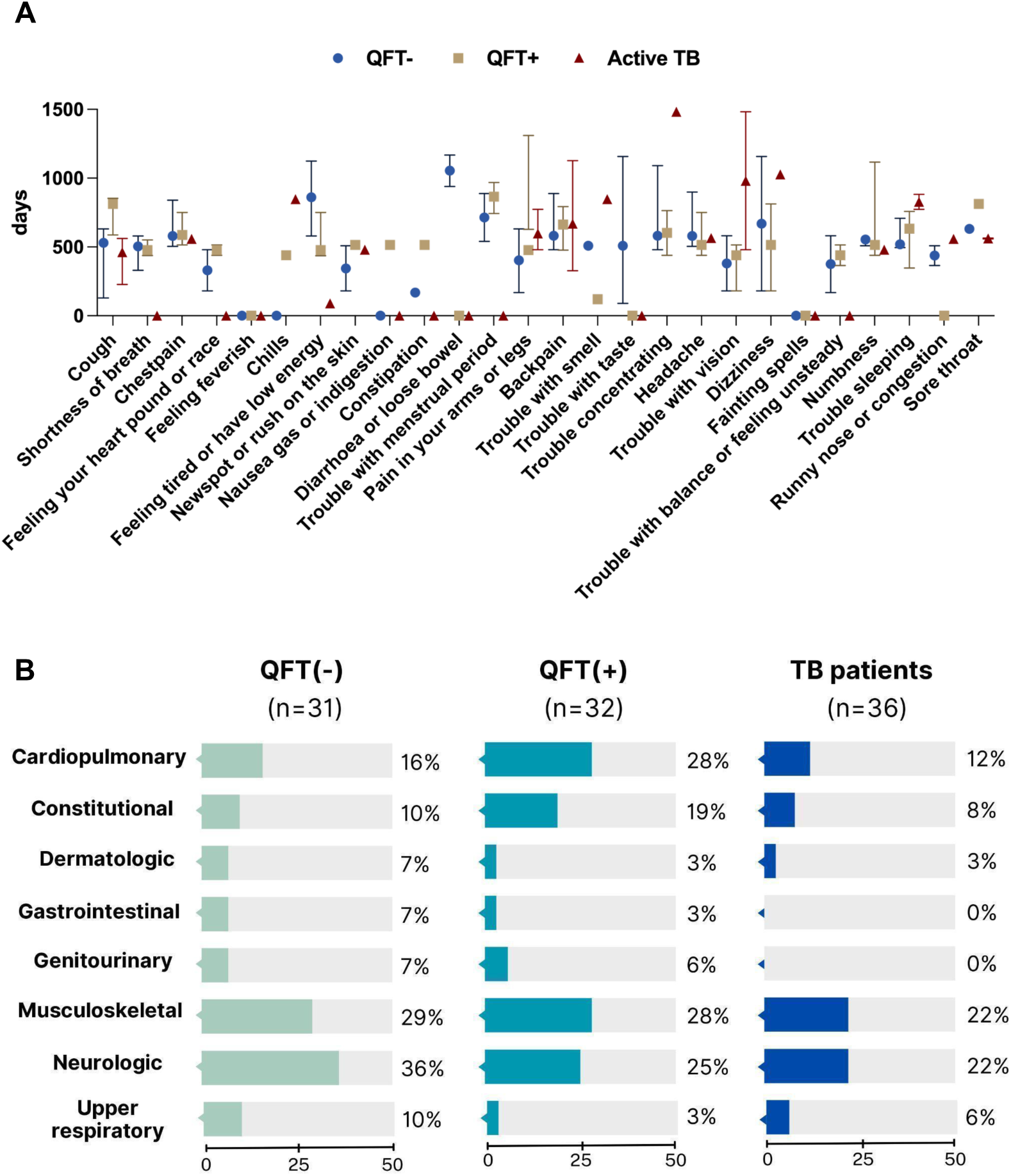
Long COVID symptoms in the Peruvian cohort. **A.** Median duration of Long COVID-19 symptoms by groups. **B.** Long COVID symptoms prevalence grouped by organ systems.

**Figure 2:**
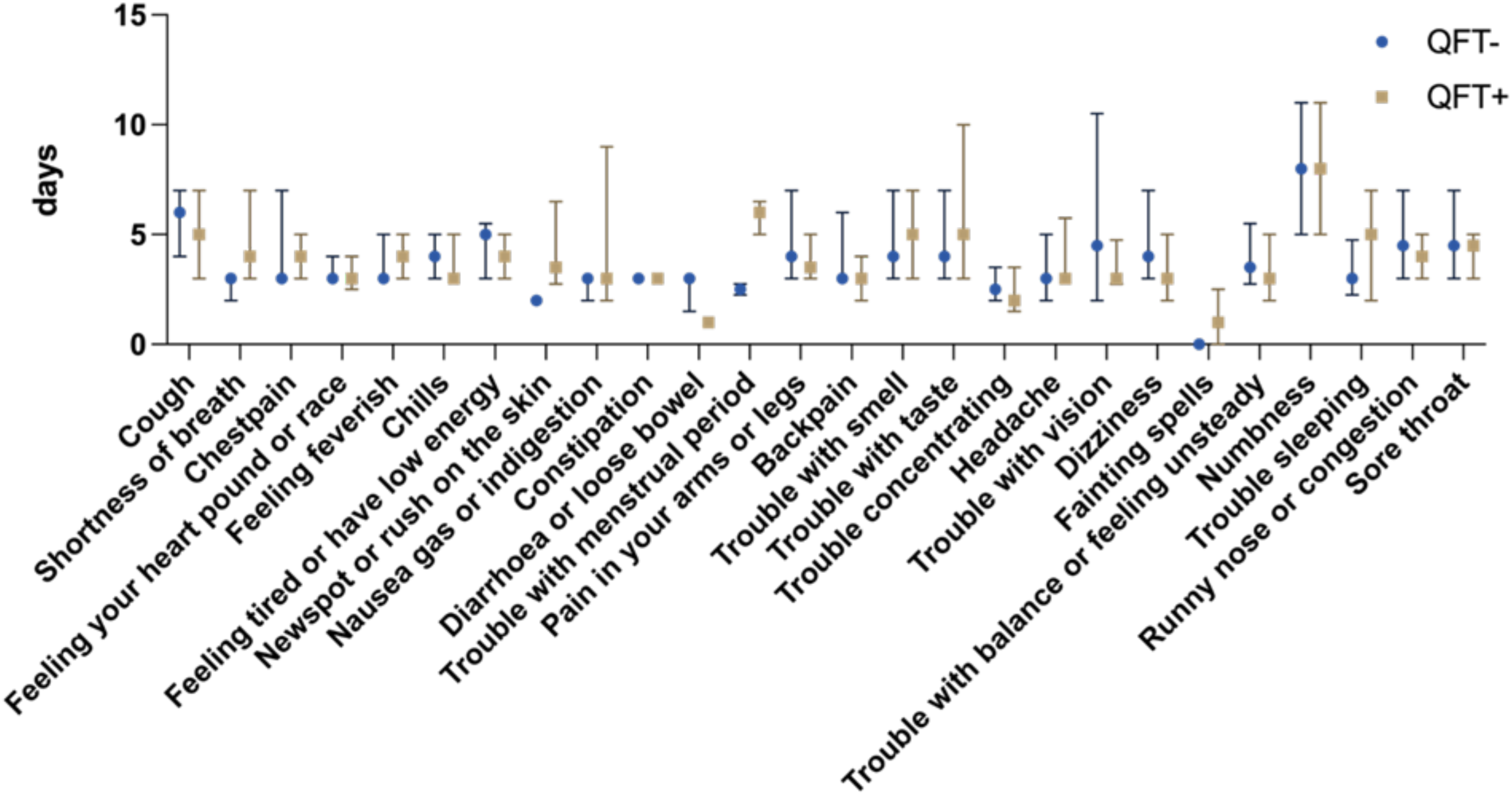
The median duration of COVID-19 symptoms by QFT+ and QFT-in Kenya.

The most prevalent groups of symptoms in all cohorts were musculoskeletal and neurological symptoms. Moreover, in the QFT+ participants, the cardiopulmonary symptoms were common (28%) (Figure 1b). While common in the acute phase, none of the participants reported fever or chills as an LC symptom (Table S1).

### Role of the Acute COVID-19 Symptoms in Predicting LC

We hypothesized that some acute COVID-19 symptoms that were prevalent at the time of SARS-CoV-2 infection were more likely to predict LC incidence. We calculated the Prevalence Ratio (PR) to test the association between acute COVID-19 symptoms and the likelihood of developing LC (Figure 3). We compared participants who experienced the symptoms for more than 10 days to the reference group who did not experience the symptoms or experienced them for less than 10 days. Adjusting by sex, age, and BMI, the symptoms more strongly associated (p<0.001) with LC were chest pain (RR [95% confidence interval (CI)]: 2.69, [1.8-4.02]), tachycardia (1.91 [1.35-2.70]), menstrual pain (2.02 [1.37-2.99]), pain in the arms or legs (2.48[1.63-3.79]), backache (3.24 [2.11-4.97]), trouble with memory (2.05 [1.39-3.02]), headache (2.14 [1.45-3.15]), trouble with vision (2.24 [1.54-3.26]), dizziness (1.82 [1.35-2.45]), and numbness (1.94 [CI 1.34-2.82]) (Figure 3). This data suggests that specific symptom types in the acute phase of COVID-19 can predict LC incidence in this group.

**Figure 3:**
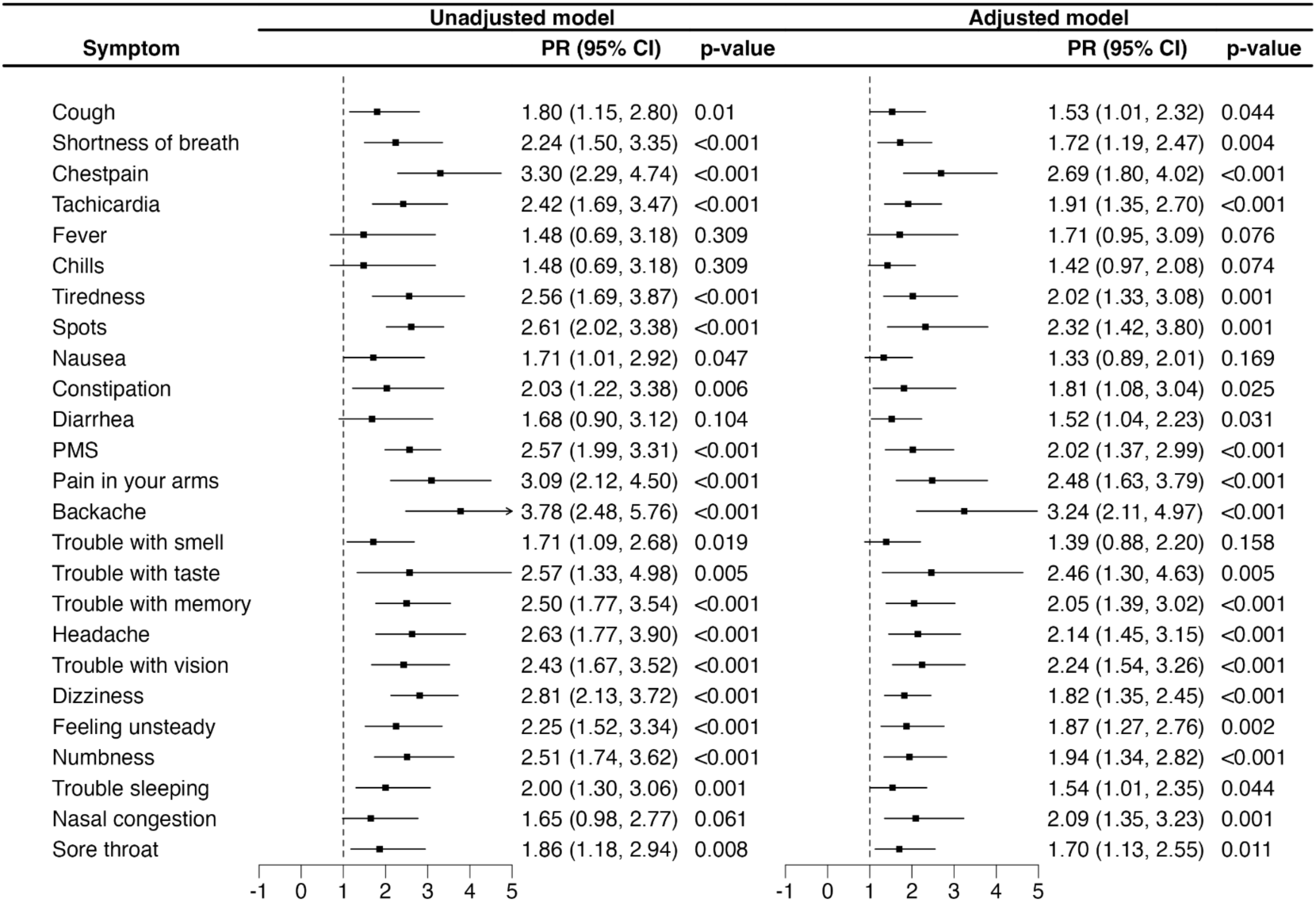
The Prevalence Ratio (PR) of Long COVID according to the persistence of the symptoms for more than 10 days in the Peruvian Cohort. The model on the right is adjusted by age, sex, and BMI.

### Influence of COVID-19 and LC on Quality of Life

We next analyzed the impact of acute COVID-19 infection and LC on self-reported quality of life and health scores in the two cohorts. We first analyzed the level of discomfort or difficulty experienced when the participants performed regular activities (Figure 4a). In the Kenyan group, participants experienced little or no difficulties performing activities before COVID-19. However, there were differences before and during the COVID-19 episode, particularly in mobility, self-care, and pain/discomfort (p<0.001), which returned to baseline levels after the acute phase of infection. We did not collect these data in the acute phase of COVID-19 in the Peruvian cohort. However, there was a significant difference between the ability to perform the same activities before and after COVID-19 (Figure 4b, p<0.01). Interestingly, depression and anxiety did not show a similar trend (p=0.289). There was no significant difference in the self-reported health scores between QFT- and QFT+ groups in the Kenyan site (Figure 4c). In the Peruvian cohort, there was a significant difference in self-reported health scores between TB groups in the pre-COVID period (p=0.01), but not during acute (p=0.98) or post-acute COVID-19 infection episodes (p=0.247).

**Figure 4:**
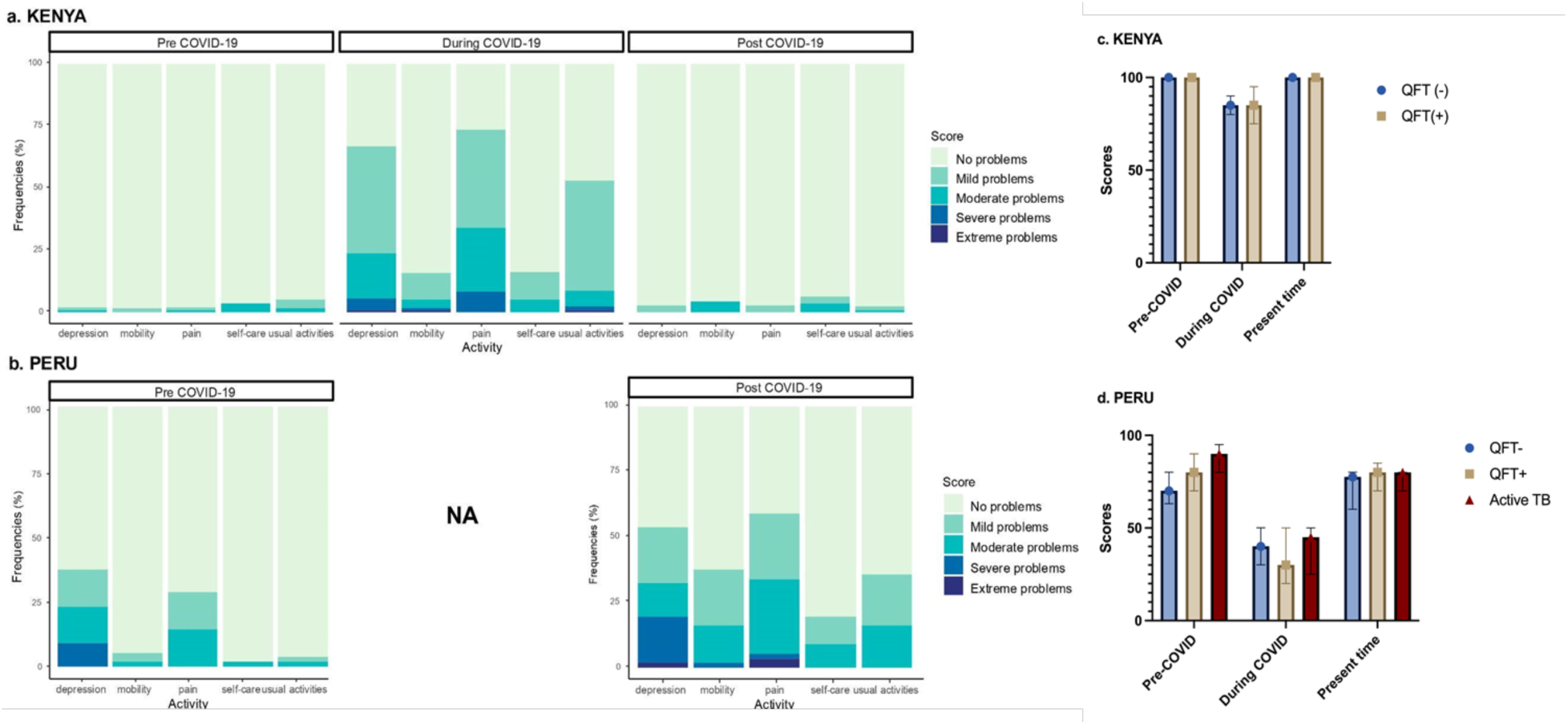
Quality of Life in both cohorts. a. Pre COVID-19, during COVID-19 and Post COVID-19 prevalence of difficulties found while performing an activity in the Kenyan Cohort. b. Pre COVID-19 and Post COVID-19 prevalence of difficulties found while performing an activity in the Peruvian Cohort. c. Self-reported health scores in the Kenyan Cohort. d. Self-reported health scores in the Peruvian Cohort.

## Discussion

LC is a major global health concern, estimated to affect tens of millions of people worldwide with symptoms like fatigue, cognitive dysfunction, and respiratory issues, lasting months after the acute phase of illness (6). The prevalence of LC in LMICs is understudied, and the impact of respiratory pathogens like *Mtb* on LC risk remains unclear (32). Here, we apply study instruments that have been used to describe the condition in the U.S. and report on the presence of LC in two cohorts in Peru and Kenya, making new observations about the presence of LC and the potential influence of *Mtb* co-infection in both settings. Our findings suggest that larger studies are needed to understand the natural history of LC in settings in LMICs, and to understand the potential interactions between SARS-CoV-2 and other pathogens, which are endemic in these regions.

We observed remarkable similarities in the persistence of LC symptoms between our Peruvian cohort (41%) and prior descriptions of LC in high-income countries (33). A meta-analysis of 421 studies estimated the pooled prevalence of LC to be around 36%, with lower rates in North America and higher rates in South America (34). This data was supported by other meta-analyses of 48 studies, which reported that the burden of LC varies depending on biological sex and geographical region, most of the studies included in this analysis were in North America (35).

Our study included people with COVID-19 history from 2020 to 2023, ranging from 4-36 months after COVID-19. A study showed that the prevalence of LC symptoms after two years of SARS-CoV-2 infection was 30%. In addition, the most prevalent symptoms were fatigue (28.0%, 95%CI 12.0-47.0), cognitive impairments (27.6%, 95%CI 12.6-45.8), and pain (8.4%, 95%CI 4.9-12.8) (36)

The most prevalent LC symptoms in our cohort, clustered by organ system, were musculoskeletal and neurological. Systematic reviews reporting LC symptoms found that fatigue and dyspnea were the most frequent, with a pooled prevalence ranging from 35 to 60% (37). Another study characterized two types of LC symptoms as acute post-COVID symptoms if they lasted less than 12 weeks and chronic post-COVID if they persisted for more than 12 weeks. This study found that the most prevalent acute post-COVID symptoms were fatigue and dyspnea (37% and 35%), and fatigue and sleep disturbance in the chronic phase (48% and 44%) (38). Collectively, these studies demonstrate the generalizable post-COVID persistence of musculoskeletal and neurological symptoms, and the need for interventions targeting these groups of symptoms.

Our study corroborated the lasting impact of COVID-19 on several quality of life measures reported in other studies. A study evaluated the cognitive functioning in people with post-COVID conditions and found higher self-reported functional impairment, a lower likelihood of full-time employment, and more severe depressive symptoms (39). In Peru, significant differences were found in mobility, self-care, pain, and daily activities, but no significant changes in depression and anxiety were noted before and after COVID-19. This could be due to other factors affecting mental health, like pre-existing diseases (39), although trends suggest an increase in depression and anxiety post-COVID (36,40). The social and economic burdens of the COVID-19 pandemic can also confound the persistence of depression and anxiety.

Several hypotheses have been proposed to explain the etiology of LC (6). Some predictors include female sex (8,41), older age (42,43), vaccination or prior infection (44,45), more severe acute disease (46) and comorbidities (47). Interestingly, a meta-analysis pointed out that although older age was predictive of LC in many studies, the pooled analysis suggested a trend in the opposite direction (48). Our Peruvian cohort was relatively young, with a median age of 34, suggesting that age was unlikely to be a major driver of LC.

In addition to age, a systematic review showed a protective role of COVID-19 vaccination against LC but recommended additional studies to validate their findings (49). Our Peruvian cohort had high COVID-19 vaccination rates, suggesting that other factors drive LC, such as comorbidities, as previously reported (50). While BMI was reported to be positively associated with LC (30), participants with TB disease in our study had lower BMI, and their contacts had higher BMI, with no difference in LC prevalence between the two groups. However, the lack of a significant difference could be due to the small sample size. Collectively, the high prevalence of LC in this young adult Peruvian cohort suggests the involvement of previously underappreciated risk factors, which require additional profiling studies.

To our surprise, none of the Kenyan participants met the criteria for LC diagnosis. According to a recent systematic review, the prevalence of LC reported in African countries ranged widely from 2% in Ghana to 86% in Egypt (51,52), suggesting heterogeneity across African populations. Our cohort of healthcare workers regularly used masks, and were thus less likely to be exposed to high SARS-CoV-2 viral loads while providing health care. Although our study did not measure the SARS-CoV-2 viral load, a study previously reported that a lower SARS-CoV-2 viral load at the time of infection was associated with a lower risk of LC (53). Other likely possibilities include recall bias, misattribution of LC symptoms to other causes, insensitivity of the instrument to measuring symptoms in the Kenyan population, cultural factors, and selection bias if people were less likely to get sick with COVID-19, since this group of healthcare workers was generally a healthy population. In addition, the stress of patient care during the pandemic may have masked the prevalence of COVID-19 symptoms.

To our knowledge, our study is among the first ones to report LC in a population highly exposed to *Mtb* in an LMIC setting. The impact of TB disease or exposure on COVID-19 outcomes is underexplored. When the pandemic started, it was hypothesized that bacille Calmette-Guérin (BCG) vaccination could provide protection against severe COVID-19, which was the basis of several clinical trials (54), but the results ultimately demonstrated no efficacy (55). Both Peru and Kenya have universal BCG vaccination guidelines, thus, BCG vaccination rates are unlikely to drive cohort-specific differences in LC prevalence. Recent studies suggest that the coexistence of *Mtb* and viruses in the lungs may increase the risk of active TB by modulating the immune system (reviewed in 56). Clinical evidence suggests that COVID-19 may predispose patients to TB disease or reactivation of latent *Mtb* infection through severe depletion and dysfunction of T-cells and uncontrolled production of pro-inflammatory cytokines (57). TB disease was shown to be associated with severe COVID-19, and *Mtb/*SARS-CoV-2 co-infection may lead to exacerbated disease (57). In our Peruvian cohort, no differences in LC occurrence were found amongst participants with active TB compared to contacts, but LC prevalence was high in all groups.

Our study has several limitations. First, our sample size is relatively small, which may limit the generalizability of our findings in both populations. Second, LC diagnosis relies on self-reports of various symptoms, which could introduce biases such as recall or social desirability bias among healthcare workers. Third, our cohorts represent convenience sampling of individuals in both settings. The decision to participate could be influenced by the presence or absence of LC symptoms in ways that are not predictable. Finally, our analysis was limited to clinical symptomatology; further work could explore whether biological drivers in our cohort are similar to other cohorts using the same instrument, such as LIINC in the U.S. (21). Larger studies could therefore define nuanced biological influences of *Mtb* co-infection in more controlled cohorts.

## Conclusion

We identified a similar proportion and similar clinical phenotypes of LC in a Peruvian cohort but did not identify LC in a cohort of Kenyan health workers. TB status was not associated with the presence of LC. Our findings underscore the need for follow-up strategies for COVID-19 patients and larger studies to explore the impact of TB on the persistence of COVID-19 symptoms.

## Data Availability

All data produced in the present study are available upon reasonable request to the authors

## Acknowledgments

S.S., S.L., C.L.A., and J.G. conceived the study idea and design. A.O. and A.C. were the study coordinators in Kenya and Peru, respectively. C.S., A.O., P.W., B.K. recruited participants and collected samples in Kenya. A.C., L.S. and D.P., recruited participants and collected samples in Peru. L.R., C.S., A.O. and A.C. performed data analysis, statistical evaluations and generated figures. J.R. and A.R. managed the coordination between the study sites. A.O. and A.C. wrote the initial draft of the manuscript. S.S, C.L.A., T.H. and M.P. reviewed and revised the manuscript critically. All authors approved the manuscript’s final version. Grant support was provided by the NIH (1R01AI169534) to J.K. and the Nina Ireland Program for Lung Health to S.S. We thank Dorcas Ihuthia and Mary Gitonga for laboratory analysis assistance, Mr. Geoffrey Okallo (CRDR, KEMRI) for data management assistance, Bryson Jumba for participant recruitment, and Mbagathi Hospital participants and recruitment site staff, Centre for Respiratory Diseases Research (CRDR), and KEMRI laboratory staff for technical assistance. In Peru, we thank the Maria Auxiliadora Hospital for assistance during the recruitment, especially Dr. Maria Paredes, Ministry of Health (MINSA) community centers for their support in the recruitment, especially Dr. Francisco Aguilar, Dr. Ronald Salazar, Dr. Carmen Melendez, and Mr. Juan Orellana.

