## Supplementary Table 1 for "Prevalence of Long COVID in *Mycobacterium tuberculosis-*exposed Groups in Peru and Kenya"

**Supplementary Data**

Table S1. Acute COVID-19 symptoms reported by the participants in both cohorts.

|  | | **Peru Cohort** | | | | **Kenya Cohort** | | |
| --- | --- | --- | --- | --- | --- | --- | --- | --- |
| **Symptoms** | | **Total, n=99** | **QFT(-)*, n=31** | **QFT(+)*, n=32** | **Active TB*, n=36** | **Total, n=202** | **QFT(-)*, n = 94** | **QFT(+)*, n = 108** |
|  |  | n (%) | n (%) | n (%) | n (%) | n (%) | n (%) | n (%) |
| Cardiopulmonary | Cough | 62 (62.6) | 20 (64.5) | 25 (78.1) | 17 (47.2) | 95 (44.0) | 40 (42.6) | 46 (42.6) |
|  | Shortness of breath | 41 (41.4) | 14 (45.2) | 13 (40.6) | 14 (38.9) | 53 (24.5) | 20 (21.3) | 29 (26.9) |
|  | Chestpain | 38 (38.4) | 14 (45.2) | 14 (43.8) | 10 (27.8) | 59 (27.3) | 22 (23.4) | 32 (29.6) |
|  | Feeling your heart pound or race | 35 (35.4) | 12 (38.7) | 13 (40.6) | 10 (27.8) | 30 (13.9) | 14 (14.9) | 14 (13.0) |
| Constitutional | Feeling feverish | 69 (69.7) | 20 (64.5) | 23 (71.9) | 26 (72.2) | 119 (55.1) | 53 (56.4) | 58 (53.7) |
|  | Chills | 56 (56.6) | 16 (51.6) | 19 (59.4) | 21 (58.3) | 109 (50.5) | 49 (52.1) | 53 (49.1) |
|  | Feeling tired or have low energy | 73 (73.7) | 24 (77.4) | 21 (65.6) | 28 (77.8) | 130 (60.2) | 63 (67.0) | 58 (53.7) |
| Dermatologic | Newspot or rush on the skin | 7 (7.1) | 2 (6.5) | 3 (9.4) | 2 (5.6) | 5 (2.3) | 2 (2.1) | 3 (2.8) |
| Gastrointestinal | Stomach pain | 22 (22.2) | 6 (19.4) | 5 (15.6) | 11 (30.6) | 4 (1.9) | 2 (2.1) | 2 (1.9) |
|  | Nausea gas or indigestion | 22 (22.2) | 6 (19.4) | 10 (31.3) | 6 (16.7) | 19 (8.8) | 12 (12.8) | 7 (6.5) |
|  | Constipation | 10 (10.1) | 4 (12.9) | 3 (9.4) | 3 (8.3) | 3 (1.4) | 1 (1.1) | 2 (1.9) |
|  | Vomiting | 12 (12.1) | 2 (6.5) | 6 (18.8) | 4 (11.1) | 3 (1.4) | 1 (1.1) | 2 (1.9) |
|  | Diarrhea or loose bowel | 28 (28.3) | 13 (41.9) | 7 (21.9) | 8 (22.2) | 8 (3.7) | 7 (7.4) | 1 (0.9) |
| Genitourinary | Trouble with menstrual period | 6 (6.1) | 2 (6.5) | 4 (12.5) | 0 (0.0) | 4 (1.9) | 2 (2.1) | 2 (1.9) |
| Musculoskeletal | Pain in your arms or legs | 39 (39.4) | 14 (45.2) | 13 (40.6) | 12 (33.3) | 65 (35.1) | 28 (29.8) | 33 (30.6) |
|  | Backpain | 56 (56.6) | 18 (58.1) | 19 (59.4) | 19 (52.8) | 23 (10.6) | 11 (11.7) | 11 (10.2) |
| Neurologic | Trouble with smell | 56 (56.6) | 17 (54.8) | 19 (59.4) | 20 (55.6) | 71 (32.9) | 37 (39.4) | 33 (30.6) |
|  | Trouble with taste | 64 (64.6) | 20 (64.5) | 20 (62.5) | 24 (66.7) | 80 (37.0) | 38 (40.4) | 36 (33.3) |
|  | Trouble concentrating, trouble with memory, thinking | 26 (26.3) | 7 (22.6) | 12 (37.5) | 7 (19.4) | 7 (3.2) | 4 (4.3) | 3 (2.8) |
|  | Headache | 75 (75.8) | 20 (64.5) | 26 (81.3) | 29 (80.6) | 93 (43.1) | 41 (43.6) | 44 (40.7) |
|  | Trouble with vision | 27 (27.3) | 8 (25.8) | 8 (25.0) | 11 (30.6) | 7 (3.2) | 3 (3.2) | 4 (3.7) |
|  | Loss of appetite | 52 (52.5) | 16 (51.6) | 19 (59.4) | 17 (47.2) | 62 (28.7) | 29 (30.9) | 31 (28.7) |
|  | Dizziness | 30 (30.3) | 9 (29.0) | 13 (40.6) | 8 (22.2) | 37 (17.1) | 19 (20.2) | 17 (15.7) |
|  | Fainting spells | 1 (1.0) | 0 (0.0) | 0 (0.0) | 1 (2.8) | 2 (0.9) | 0 (0.0) | 2 (1.9) |
|  | Trouble with balance or feeling unsteady | 28 (28.3) | 10 (32.3) | 11 (34.4) | 7 (19.4) | 9 (4.2) | 4 (4.3) | 5 (4.6) |
|  | Numbness, tingling, or “pins and needles” in your arms or legs". | 30 (30.3) | 12 (38.7) | 10 (31.3) | 8 (22.2) | 4 (1.9) | 2 (2.1) | 2 (1.9) |
|  | Trouble sleeping | 42 (42.4) | 13 (41.9) | 18 (56.3) | 11 (30.6) | 33 (15.3) | 14 (14.9) | 18 (16.7) |
| Upper respiratory | Runny nose or congestion | 50 (50.5) | 17 (54.8) | 17 (53.1) | 16 (44.4) | 78 (36.1) | 35 (37.2) | 37 (34.5) |
|  | Sore throat | 60 (60.6) | 20 (64.5) | 22 (68.8) | 18 (50.0) | 90 (41.7) | 37 (39.4) | 46 (42.6) |

**Active TB: includes TB patients at different points in antibiotic treatment; QFT (-): TB household contact with negative Quantiferon test; QFT (+): TB household contact with positive QuantiFERON TB Gold test
